# CyclOps: Cyclical development towards operationalizing ML models for health

**DOI:** 10.1101/2022.12.02.22283021

**Authors:** Amrit Krishnan, Vallijah Subasri, Kaden McKeen, Ali Kore, Franklin Ogidi, Mahshid Alinoori, Nadim Lalani, Azra Dhalla, Amol Verma, Fahad Razak, Deval Pandya, Elham Dolatabadi

## Abstract

Open source software that enable research and development of machine learning (ML) models for clinical use cases are fragmented, poorly maintained and fall short in functionality. CyclOps is a software framework designed to address this gap and help accelerate the development of ML models for health. In this paper, we describe the architecture, APIs and implementation details of CyclOps, while providing benchmarks on example clinical use cases. We emphasize that CyclOps is developed to be researcher friendly, while providing APIs for building end-to-end pipelines for model development as well as deployment. We adopt software engineering and ML operations (MLOps) best practices, while providing support for handling large volumes of health data. The design of the framework is centered around the notion of iterative and cyclical development of the overall ML system, which consists of data, model development and monitoring pipelines. The core `CyclOps` package can be installed through the Python Package Index (PyPI) and the source code is available at https://github.com/VectorInstitute/cyclops.

## 1 Introduction

Healthcare continues to evolve with unforeseen challenges, and as it becomes more complex, there is a need for solutions to improve the quality of clinical care. Machine learning hopes to address some of these challenges, by increasing clinician capacity and improving efficiency such that they can provide better care for patients. While we expect the number of health ML models being developed to rise over time, adoption of these models in clinical practice appears to lag, with limited use in hospitals despite recent efforts to make health data repositories such as MIMIC [8] [6], U.K [19] and Japan [12] Biobanks, eICU [16], I2b2 [11], ChexPert [4], NIH open to the public. Major barriers to adoption include poorly designed ML workflows, namely due to a lack of tools for rigorous model development and evaluation, as well as tools for integrating MLOps into clinical workflows.

Managing tasks in an ad-hoc manner during ML development makes the execution of steps error-prone, difficult, time-consuming, and also prevents developers from building sophisticated ML models. This raises barriers towards improving and extending development workflows, leads to unmanageable code and prevents building scalable systems. Hence, software that is easy to modify and extend, while remaining modular and well tested is necessary for adoption of AI systems in healthcare. Figure 1 shows the cyclical workflow of developing and operationalizing an ML model. This workflow, while typical for ML system development, needs to be designed and developed specific to the needs and complexities of healthcare.

**Figure 1:**
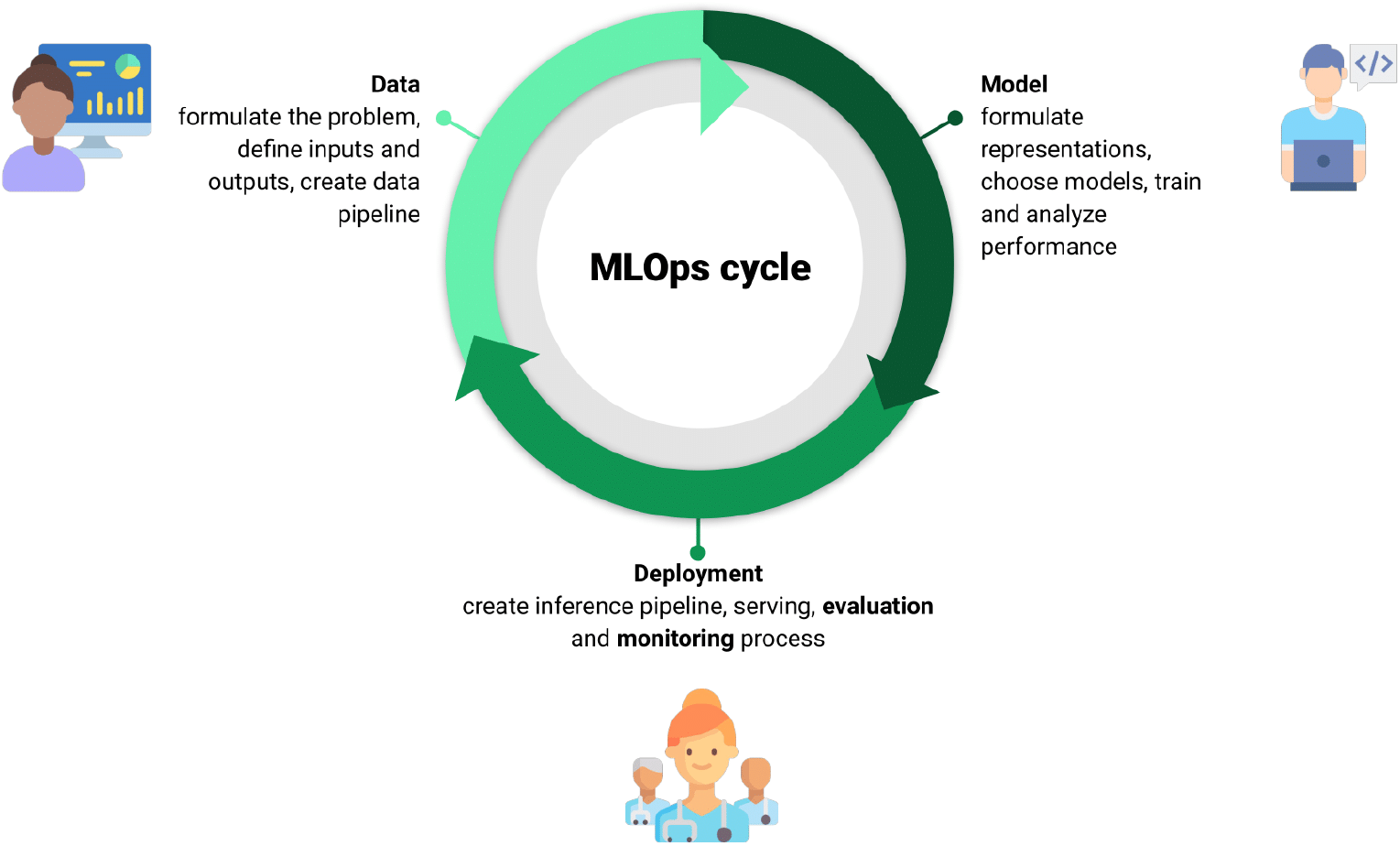
The cyclical nature of developing and operationalizing an ML model. We view this typical workflow through the unique lens of healthcare for designing CyclOps.

A wide variety of tools and engineering pipelines have recently been developed in the context of health ML, but were designed for specific use cases and patient populations, and applied within the narrow context in which they were developed. Moreover, existing approaches have been developed for research use cases and lack software engineering rigour such as testing, documentation and continuous integration. In general, they fail to scale for large volumes of data and do not generalize well across use cases.

CyclOps was built to address these shortcomings and overall provide support for ventures at the intersection of ML and health. We invite open-source collaboration to drive the building of use cases and tools using CyclOps. We hope to support and maintain the software as it grows and gets refactored towards improved usability and functionality.

We have evaluated CyclOps and demonstrated its early usability and performance on two distinct use cases. The first use case is built on clinical electronic health record (EHR) data for general internal medicine patient populations where the goal of the ML models is to predict the risk of mortality. We have also demonstrated a use case using imaging data, and hence showcase the extensibility of the framework to multimodal data.

## 2 Related Work

There have been multiple pioneering efforts to address the issue of reproducibility, specifically in the context of machine learning research using medical datasets. Most notably, the `mimic-code` repository, a toolkit that processes the Medical Information Mart for Intensive Care (MIMIC) into an accessible relational database [7], has enabled researchers to easily set up the database and extract data towards their research use cases.

`MIMIC-Extract` [22], is a popular open-source toolkit for transforming raw electronic health record (EHR) data from MIMIC-III into DataFrames that are directly usable for ML modelling. This toolkit provides standardized data processing functions, including unit conversion, outlier detection, and imputation functions to account for missingness in medical time-series data. Similarly, `mimic3-benchmarks` [3] is a popular open-source benchmark which showcases predictive models in order to allow researchers to produce comparable and reproducible results on the MIMIC-III dataset. The benchmark provides baselines on four tasks, namely in-hospital mortality prediction, decompensation prediction, length of stay prediction and phenotype classification. Other tools and benchmarks on the MIMIC-IV [6] [2] dataset, eICU-CRD [16], HiRID-ICU-Benchmark [24], AmsterdamUMCdb [20] also provide processing pipelines and baselines for researchers. `ATLAS` [13] is built by the Observational Health Data Sciences and Informatics (OHDSI) organization and can be used to conduct analyses on datasets that are available in the Observational Medical Outcomes Partnership (OMOP) format. `PyHealth` [25] is a Python library that offers preprocessing across various EHR datasets and modelling for multiple clinical prediction tasks. Moreover, these tools fail to offer capabilities to evaluate model robustness to data shifts and existing tools for monitoring of clinical ML models like `CheXstray` [18] focus on specific domains and offer limited functionality.

A seminal work to showcase the importance of pipelines and modular implementations is `Clairvoiyance` [5]. The toolkit highlights the challenges of engineering quality software to process medical time series data. The authors of `Clairvoiyance` also show that a unified approach where the data processing and modelling along with providing functions to obtain model calibration, interpretation scores, evaluation and other useful analyses is needed for clinical ML problems. `Clairvoiyance` highlights the need for a one-stop framework for clinical ML implementation, however it was built as a proof-of-concept research toolkit with limited scope and applications.

## 3 Design Principles

- **Evaluation and monitoring centric** CyclOps is built to enable the rigorous evaluation and monitoring of clinical ML models. Each component is designed to enable slicing of data such that the evaluation of models across different attributes can be easily achieved. Furthermore, the APIs provided to enable evaluation and monitoring are extensible and easy-to-use.
- **Pythonic** Python is highly popular in the machine learning space, supporting many packages and tools for data processing as well as ML modelling, research, and analysis. Hence we have built the components of CyclOps using Python, and have provided APIs in Python. Under the hood, we use Pandas and NumPy libraries extensively for data processing. The interfaces are built to avoid repeated work and greatly simplify data extraction, processing and modelling workflows.
- **Researcher friendly** By providing easy-to-use Python API functions, we aim to support researchers to easily extract, process and analyze EHR data. Furthermore, the APIs provided are highly customizable and can be used to run experiments with different parameters.
- **Modular and supports pipelines** Due to the complex nature of tasks in the ML development workflow, especially for workloads involved in processing large volumes of tabular, time-series and other clinical data, CyclOps supports the building of pipelines such that these tasks can be modularized and tested. This makes it easier to debug and modify the framework components as they grow in complexity.

## 4 Application Programming Interfaces

### 4.1 Query API

The first stage of any machine learning pipeline is data extraction. Data scientists often refer to this process as Extract, Transform, Load (ETL), where data in a raw format is extracted from a database, transformed to a more clean and structured format, and loaded into a new database. Health data, and especially EHR data, is often stored in relational databases, where programs written in the Structured Query Language (SQL) are typically used to perform ETL on said databases. However, SQL code can be hard to understand and maintain, serving as a distraction and limiting the extensibility of data pipelines.

To address this problem, we introduce a query API implemented in Python. As shown in figure 2, the query API consists of low-level functions that can be used to perform common operations on the database. These functions use the SQLAlchemy toolkit which implements an Object Relational Mapper (ORM), that can be used to query relational databases. The output of the query API are Pandas DataFrames, a common representation used for static and temporal (time-series) data.

**Figure 2:**
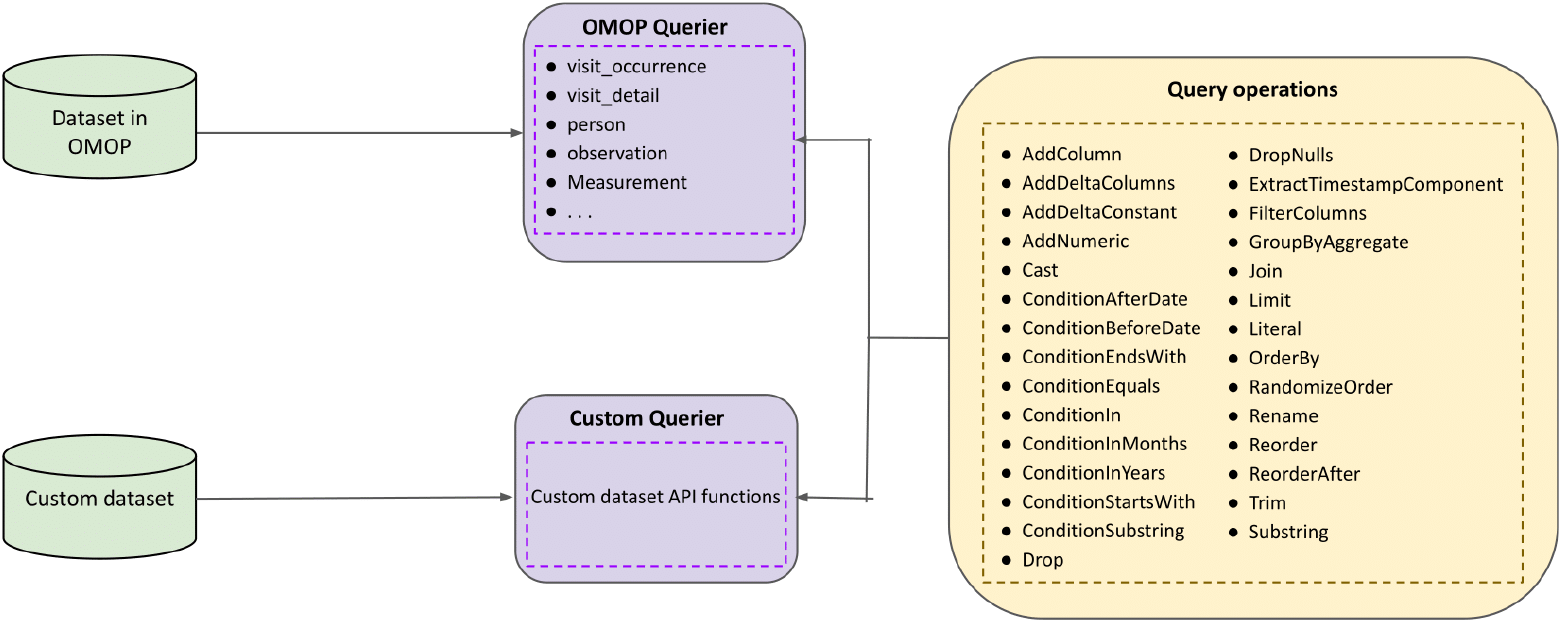
The Query API can be used to query EHR databases. The querying operations are modules implemented using the functional API, which in turn uses the SQLAlchemy toolkit. These modules can be combined sequentially to create the set of operations to apply on the database.

Operations such as casting columns to specific data types, adding a new column, joins, re-ordering, and so forth, can now be used as modules and combined sequentially. Hence, we can use these modules to create a set of high-level API functions to query EHR databases. As an example shown in code snippet 1, we can create functions that accept various arguments used to perform operations on the database. To process these arguments, we use a Query Argument Placeholder (QAP) class, which wraps around the provided arguments, which is then used to optionally apply querying operations provided the necessary arguments are specified.

The query API enables the creation of easy-to-use API functions to query EHR databases. We provide API functions to query the popular MIMIC-IV [6] database, with plans to support other popular research datasets. As shown in example code snippet 2, we can use these functions to easily extract a cohort of interest towards a use case.

We also provide high-level API functions to query datasets that are available in the OMOP Common Data Model (CDM) [13]. The adoption of a common data model for EHR data has several advantages, and several health institutions have successfully converted their data to OMOP or are in the process of doing so. Hence, our default support extends to OMOP databases. We do provide support for developers working on arbitrary databases, where the query API can still be applied after writing some wrapper code - for which there are tutorials and examples.

**Listing 1:**
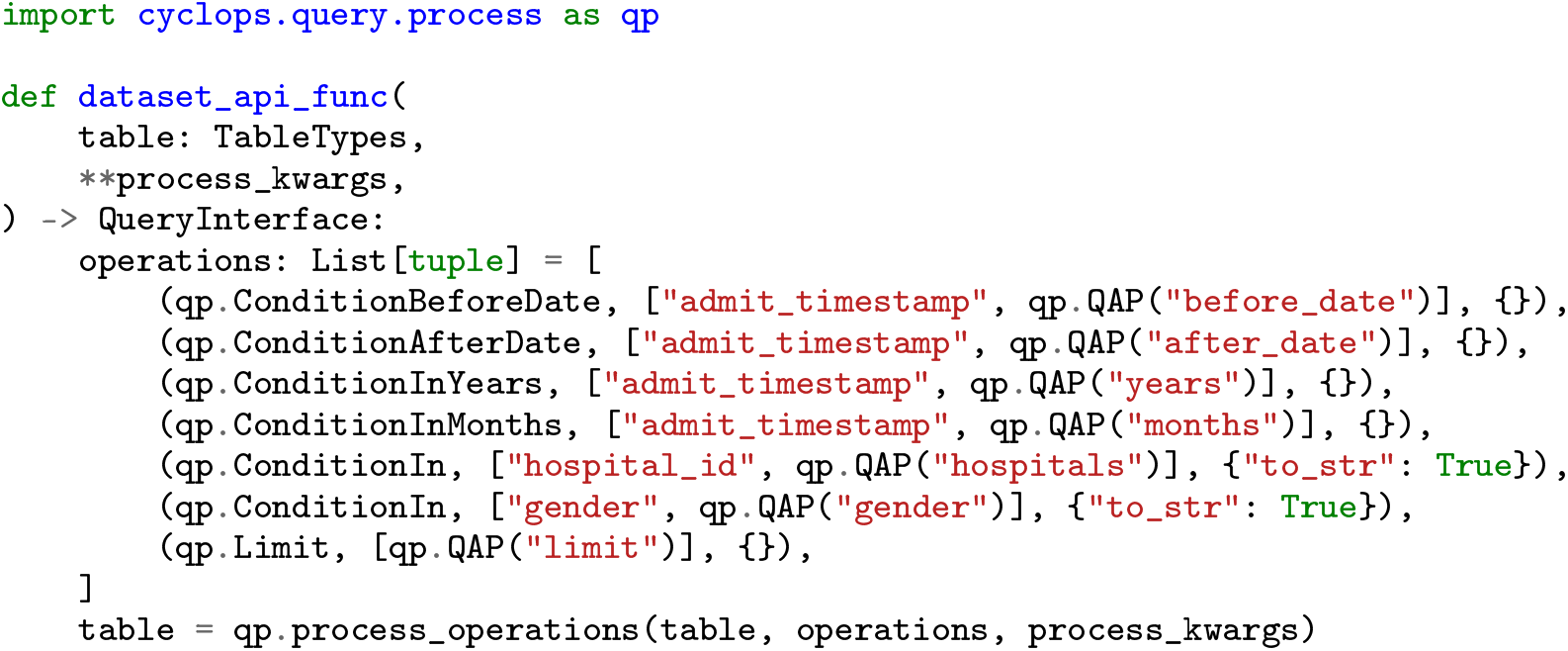
An example of a high-level API function to query a dataset. In this example, the table consists of patient administrative data, and hence arguments to the function can be used to filter the data based on admission timestamps, hospital and gender.

**Listing 2:**
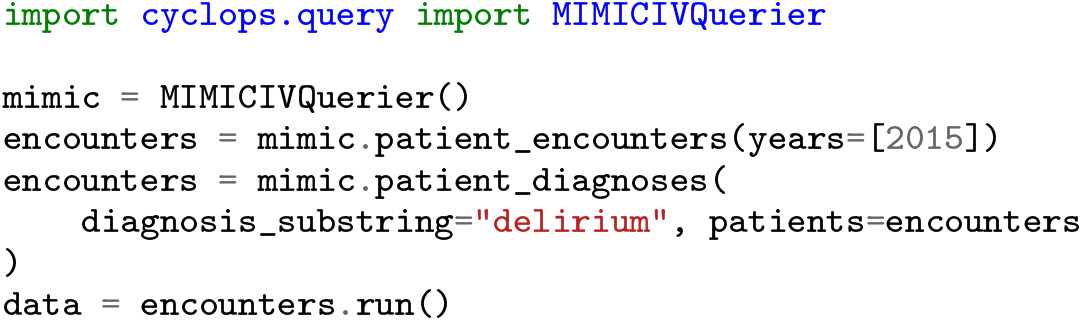
An example of the usage of the query API to extract data from the popular MIMIC-IV database. In this example, we filter patient encounters from the year 2015, and further filter those that had a diagnoses associated with delirium.

### 4.2 Process API

Once data is extracted to create a subset of interest, it needs to be processed towards creating useful representations for ML models. These features are structured for use in machine learning, for example through vectorization. NumPy arrays and PyTorch tensors are examples of vectorized data containers that are used as input to ML model implementations. Transforming raw health data into these vector representations is a multi-step process, and we provide a process API which supports:

- **Cleaning**: Formatting values, adjusting data types, unit conversion, checking and fixing issues with raw data.
- **Aggregation**: Grouping infrequent and unevenly spaced temporal measures into buckets, e.g., for aggregating lab measurements into fixed time-interval buckets, or grouping values by patient ID.
- **Normalization**: Normalizing values into standard ranges or using statistics of the data.
- **Imputation**: Handling missing values in the data. For time-series clinical data, ‘missingness’ contains valuable contextual information about the patient’s health and needs to be handled carefully. The imputing step fills missing values using various configurable approaches including mean, median and forward, and/or backward filling for time-series data.
- **Slicing**: Filtering the data based on conditions applied to columns. For example, slice the data by a column that contains age information to obtain a subset with all patients above the age of 50.
- **Dataset splitting**: Splitting the data into different subsets, for example, training, validation and testing subsets, as well as supporting cross-validation procedures.
- **Vectorization**: Converting patient representations into vectors which are ready to be used as inputs to ML models. This format typically consists of multi-dimensional arrays over which operations are more efficient.
- **Health-specific processing**: Processing steps specific to health data, such as grouping diagnosis codes (ICD-10 codes) into disease trajectories to create features. ICD-10 codes tend to be sparse and granular, and hence grouping them offers more useful representations for modelling.

As shown in Figure 3, the process API provides functionality to parse a raw DataFrame into an intermediate format to represent features, using data containers for them. These intermediate containers hold the data as Pandas DataFrames, while providing useful methods to create feature sets for analysis and modelling. There are two containers `TabularFeatures` and `TemporalFeatures`, which inherit a base `Features` class. These containers hold the extracted data, the features of interest as specified by the user and metadata information about the features, such as the type and whether it may be a target variable. If the feature is an indicator variable, the metadata also has information about its parent variable. The container automatically attempts to detect feature types which can be string, numeric, binary, or categorical indicators (dummy variables).

**Figure 3:**
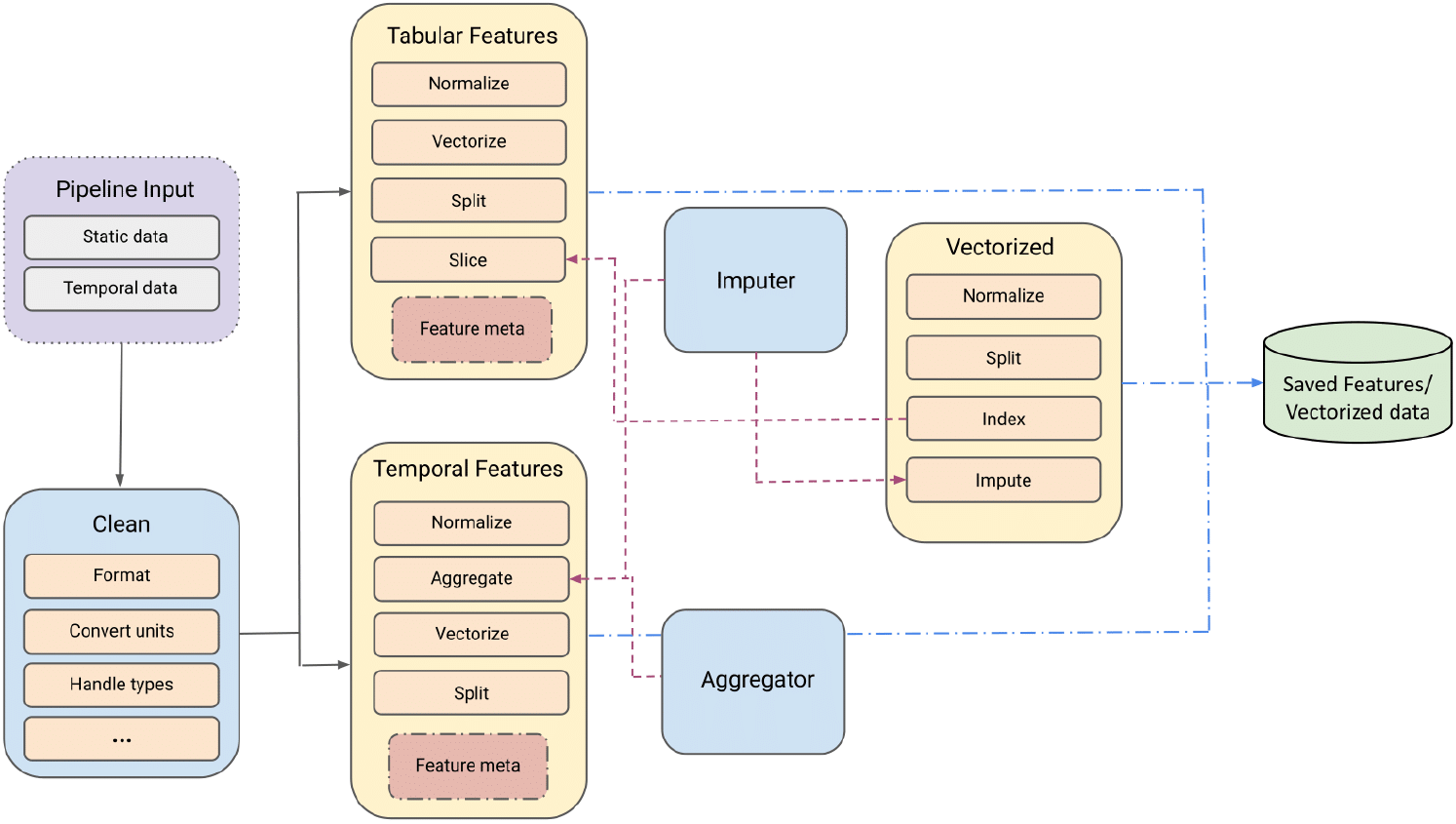
The Process API supports various processing steps towards transforming raw EHR data into features for ML models. The `Features` and `Vectorized` data containers are primarily used to featurize and vectorize the data respectively.

**Figure 4:**
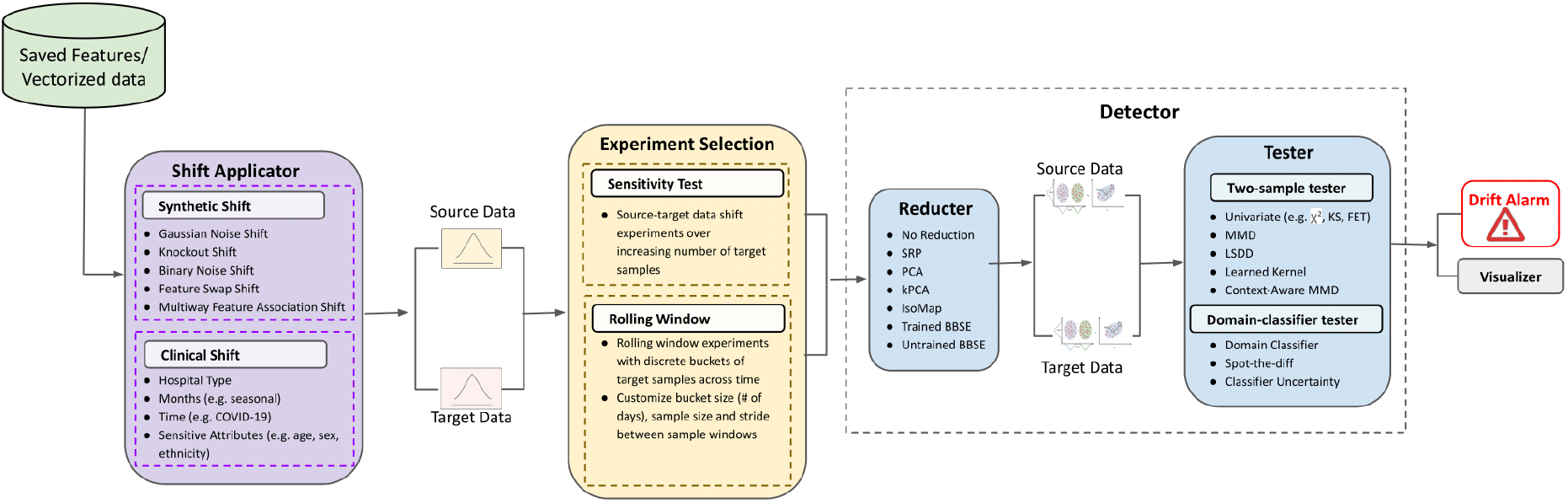
The Monitor API supports shift application, experiment selection and drift detection.

The user can also force a feature to be of a certain type, thus allowing flexibility. Furthermore, the containers support useful methods to `normalize`, `slice`, `split` and `vectorize` the features data. The `TemporalFeatures` container has an additional `aggregate` method that aggregates the time-series data. We provide an additional `Vectorized` data container for storing vectorized data. This container holds the data as NumPy arrays, and also contains useful methods to `normalize` and `split` the data. Example code snippets 3 and 4 show how to instantiate the feature containers and call methods to aggregate and vectorize the data.

Model implementations that use the scikit-learn [15] and PyTorch [14] frameworks interface well with NumPy arrays. We provide a models library that contains reference implementations of several commonly used ML models for clinical data, and showcase examples that ingest data from the `Vectorized` data containers.

### 4.3 Monitor API

As the barrier to scaling clinical AI systems, shifts from model development to deployment, there is a growing need for continuous monitoring and updating in order to understand when a model is likely to output erroneous predictions due to distributional shifts. The monitor API allows for the detection of data drift prior to model failure in order to inform end-users and trigger retraining and updating procedures. It provides support for rigorous ML model monitoring across time, patient cohorts, location, and custom data splits to enable the safe deployment of clinical ML models. The methods use state-of-the art drift detection methods [21] and are designed to work with high dimensional data by performing dimensionality reduction followed by a two-sample or classifier-based test to identify and characterize whether there is a statistically significant distribution shift between the source and target domain [17]. In this regard, the API provides the following:

**Listing 3:**
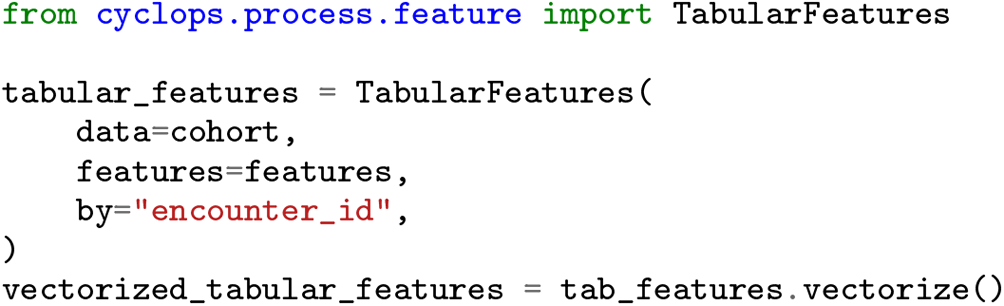
Processing of static features, using the `TabularFeatures` class.

**Listing 4:**
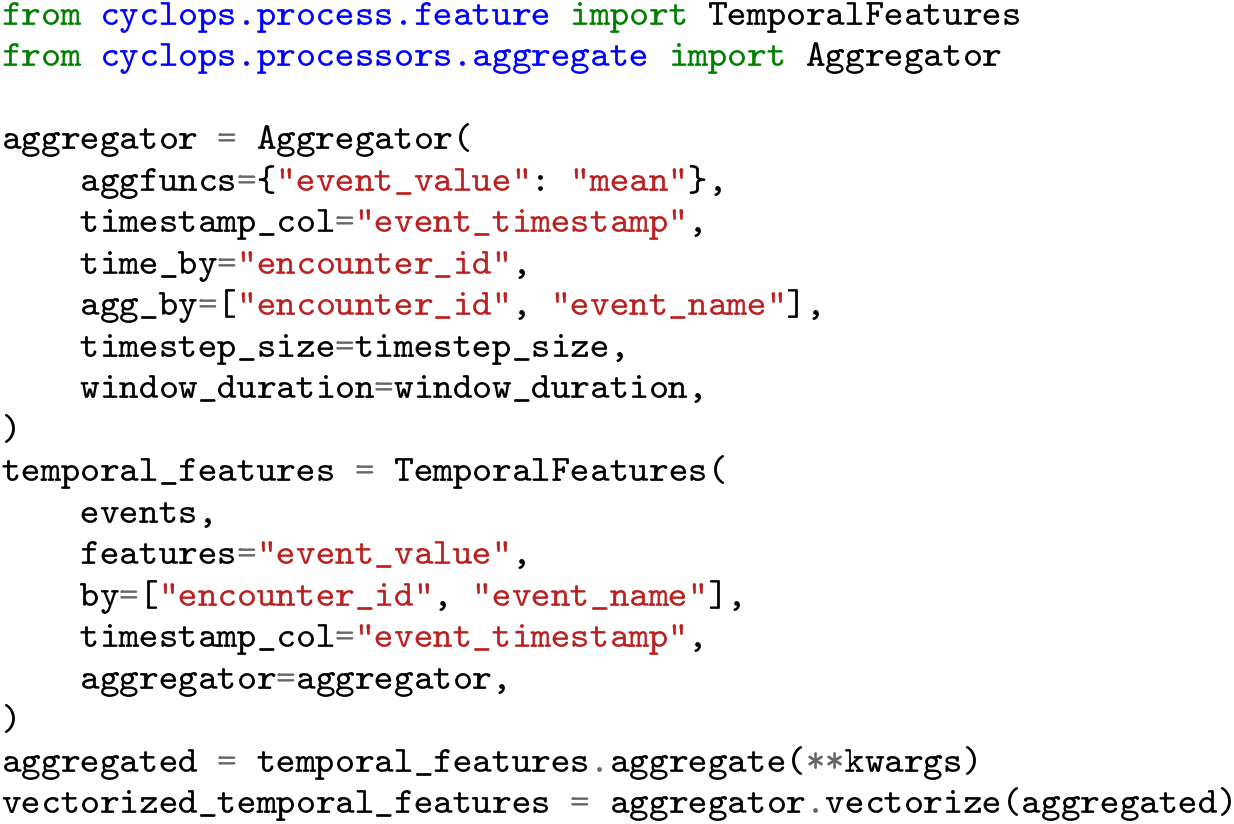
Processing of temporal features, using the `TemporalFeatures` class.

- **Clinical Shift Applicator**: Enables real clinical experiments for drift detection through splitting the data into different subsets across time, patients’ cohorts, locations, seasons, or unprecedented events such as COVID.
- **Synthetic Shift Applicator**: Perturbs a subset of data synthetically using a variety of covariates and label shifts with varying magnitudes and proportions.
- **Rolling Window Analyzer**: Evaluates the model stability and potential drift over time using discrete buckets of time.
- **Reductor**: Performs various linear and non-linear dimensionality reduction techniques to produce lower dimensional latent representations for downstream testing. The Reductor also supports dimensionality reduction using Black Box Shift Estimators (BBSE) [9] across various deep learning algorithms for EHR and imaging data.
- **Domain Classifier Tester**: Implements domain classifiers to identify if a shift in the distribution has occurred between the latent representation of source and target domains.
- **Two-Sample Tester**: Implements univariate and multivariate two-sample statistical tests to identify if a shift in the distribution has occurred between the latent representation of source and target domains.

The monitoring API can be implemented using three main steps: (1) shift application, (2) experiment selection, and (3) drift detection. As previously mentioned, the primary goal is to detect distribution shifts in clinical data.

#### Shift Application

The first step is shift application which involves the creation of source and target datasets based on criteria and conditional logic defined by the user. Given the nature of the intended shift, the user can choose to leverage the `SyntheticShiftApplicator` or the `ClinicalShiftApplicator`. The `SyntheticShiftApplicator` adds synthetic perturbations that span a variety of covariate and label shifts with varying magnitudes and fractions of affected data to the target subset of the dataset. The `ClinicalShiftApplicator` includes conditional logic to select subsets of data based on hospital, time, patient population, and other sensitive attributes (e.g. age, race, gender) to enable real-life drift detection experiments.

**Listing 5:**
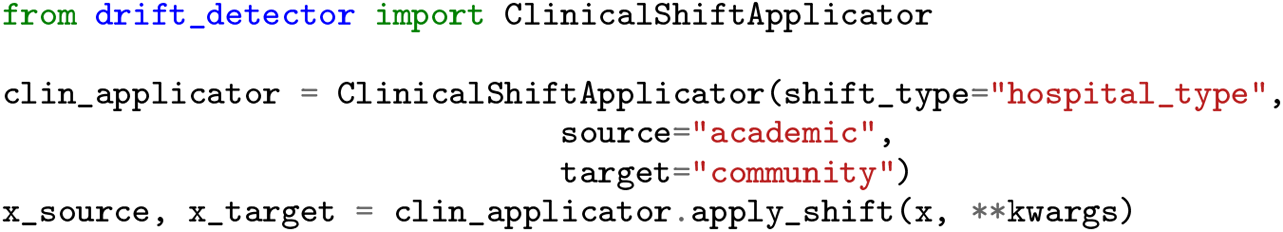
Shift Application using the `ClinicalShiftApplicator` class.

**Listing 6:**
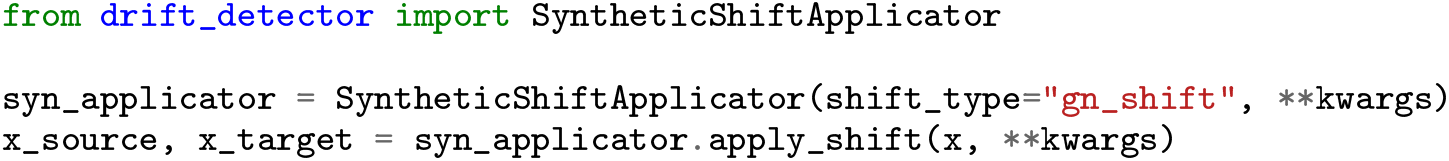
Shift Application using the `SyntheticShiftApplicator` class.

#### Experiment Selection

The next step is experiment selection with functionality to perform a sensitivity test or a rolling window experiment. The sensitivity test will setup an experiment where data distribution shift is measured between a fixed sample size from the source population and increasing sample sizes randomly sampled from the target population. The `RollingWindow` will setup an experiment where data distribution shift can be measured between i) two streams of time-varying source and target data separated by user-defined lookback period or ii) a constant set of source data used to train a model and streams of incoming time-varying target data. The size of the window, the stride length, drift threshold and number of samples to use for statistical testing can all be customized by the user.

#### Drift Detection

The final step is drift detection using the `Detector` which consists of two components: Reductor and Tester. The `Reductor` supports various techniques to reduce the dimensionality of source and targets datasets. The dimensionality reduction technique is `fit` using the source dataset and is subsequently used to `transform` the target dataset into a lower dimensional space. Following reduction, the Tester performs the two sample statistical test to detect distribution shift between the source and target datasets in the lower dimensional space. For the statistical test, users can choose between `TSTester` to perform two-sample statistical tests or `DCTester` for domain classifier-based tests. Depending on the type of experiment selected by the user, the final output of the pipeline would be either a p-value sensitivity test plot or a rolling-window plot. The p-value plot indicates the p-value for each test with increasing number of target samples along with mean and standard deviation of the p-value across multiple runs. The rolling window plot yields an array of p-values for each test across time for the rolling window scheme specified along with an array that specifies whether the p-value fell below the threshold for each time-step.

**Listing 7:**
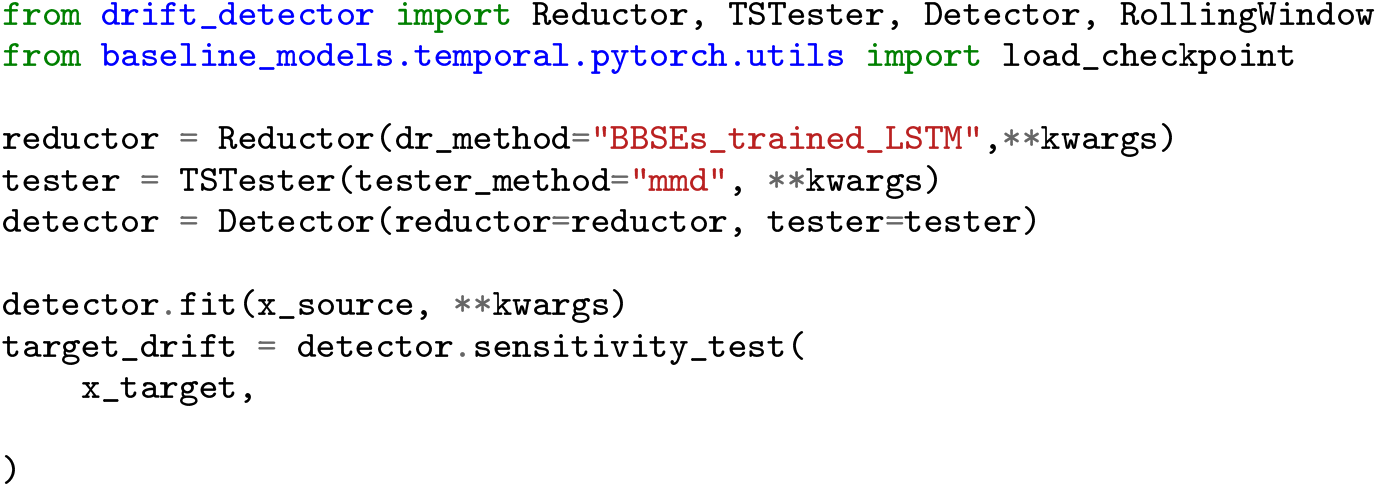
Initialize `Detector` with `Reductor` and `TSTester` and conduct sensitivity test.

**Listing 8:**
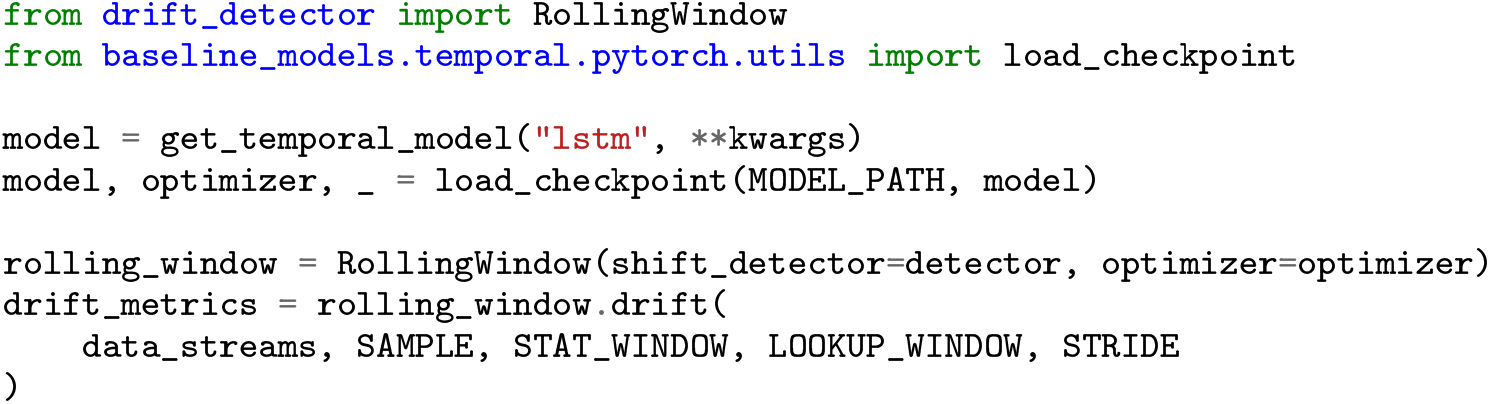
Rolling window drift experiment, using the `RollingWindow` class initialized with the `Detector`.

## 5 Proof of Concepts Experiments

We conducted a series of experiments to demonstrate how the CyclOps framework can be used for building data extraction, modeling, and drift detection pipelines. The experiments were made for various patient populations, data modalities, and outcomes.

### 5.1 General Internal Medicine Unit - GEMINI

GEMINI is a multi-center database comprising information related to patients admitted to general internal medicine units at 7 large hospitals in the Greater Toronto Area from 2010-2020. GEMINI is a relational database which includes 13 tables and 156 variables comprising administrative and clinical information. We used the GEMINI database **??** to indicate the usability of the CyclOps framework for three real-life clinical tasks: (1) in-hospital mortality risk prediction, (2) mortality decompensation prediction. We showcase three clinical use cases that leverage different combinations of input data, ML modeling, clinical outcomes, and drift detection experiments to build robust clinical AI systems using CyclOps.

#### 5.1.1 In-hospital mortality risk prediction

We used the data and drift detection pipelines of the CyclOps framework for the task of in-hospital mortality risk prediction. The data pipeline as shown in Table 1, was used to create vectorized data to be used by the ML classifier which is a Gradient Boosted Tree (GBT) to predict in hospital mortality. Given the non-temporal nature of the GBT model, the input data to the model is the combination of vectorized static data and temporal features aggregated over the first six days of patients’ stay in the GIM unit in the hospital. Only patients’ data whose length of stay (LOS) in the hospital was more than 24 hours were included in this clinical task. In total 17,365 data records were created by the data pipeline where a subset was used for GBT training and the rest for model evaluation and drift detection analysis.

**Table 1:**
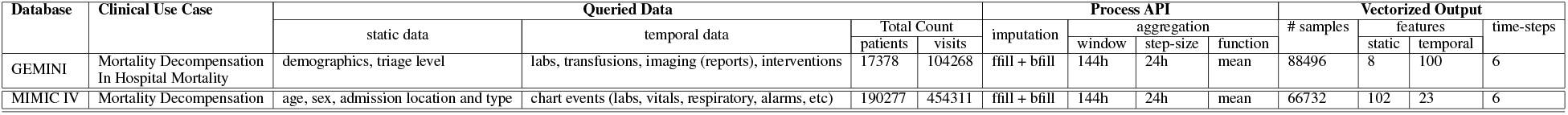
Data pipeline summary for the example use cases. The dataset sizes and parameters can be configured by researchers.

The framework’s drift detection pipeline was then leveraged to evaluate the GBT’s robustness across changing season, location, and the COVID-19 outbreak as shown in Table 2. For each drift detection experiment, the GBT model was built on source data and evaluated on out-of-distribution target data. The source and target data splits were created by the `ClinicalShiftApplicator` under the Shift Applicator module in the pipeline 4. As the results in Table 2 indicate, the performance of the model trained on community hospitals deteriorated when used to predict in-patient mortality in academic hospitals. To statistically measure data drift in a label-agnostic manner we used the CyclOps `Detector` module with a trained BBSE as the `Reductor` and MMD as the `TSTester`. Figure 5 depicts the p-value sensitivity test plot produced by the drift detection pipeline.

**Table 2:** Gradient Boosted Tree (GBT) model performance across dataset shift experiments.

|  | Splits | Clinical Applicator | # records | auROC | auPRC |
| --- | --- | --- | --- | --- | --- |
| 1 | Source | pre-COVID | 885 | 0.880 | 0.875 |
|  | Target | COVID | 1082 | 0.909 | 0.921 |
| 2 | Source | Summer | 2425 | 0.901 | 0.911 |
|  | Target | Winter | 5255 | 0.899 | 0.895 |
| 3 | Source | Community | 1076 | 0.936 | 0.971 |
|  | Target | Academic | 9830 | 0.672 | 0.574 |

**Figure 5:**
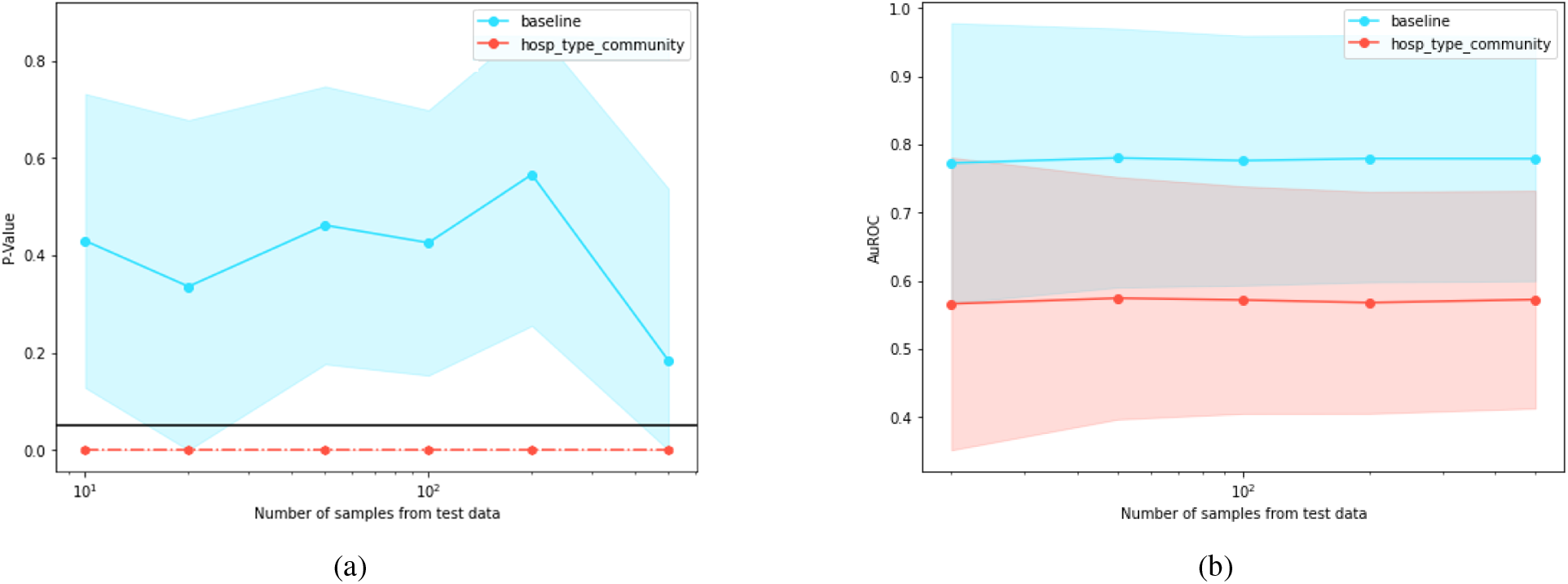
Drift detection experiments using the `Detector` for a GBT model trained on community hospitals evaluated on in-distribution data from community hospitals (blue) and out-of-distribution data from academic hospitals (red). Sensitivity across increasing number of samples from test data for a) drift p-values and b) performance (AUROC)

#### 5.1.2 Mortality Decompensation Prediction

We used CyclOps framework to build a dynamic model for predicting patient’s functional health deterioration over time. To predict functional deterioration which is risk of mortality in our case, we stepped forward through each patient’s longitudinal data, and made predictions every 24 hours for the risk of mortality within the next two weeks starting 24 hours after admission. Patient’s longitudinal data included temporal clinical measures for six consecutive days at the GIM unit. For the classification task, we used varying length LSTM model using the target replication approach [10] to predict mortality at every time step (24 hours). In addition to patient’s longitudinal clinical measures, baseline demographics as static variables are also used at every time step for the prediction task.

CyclOps data pipeline as detailed in Table 1 was used to create patients data for the prediction task. Out of 88,496 vectorized records generated by the pipeline, 46,975 was used for model development which was captured up to January (pre-COVID-19) and the rest was used for comprehensive evaluation and drift detection. The LSTM model was trained on 46,975 observations and tested on 29,738 observations with an AUROC score of 0.793 on pre-COVID-19 test data.

Following the model development, drift detection pipeline was used for rigorous evaluation of the model across time and mainly during the COVID19 time through rolling window analysis. Figure 6 illustrates the rolling window plots generated by the framework which includes trajectories across 15 months for drift: distance and p-value, and model performance: sensitivity, PPV, AUROC, and AUPRC. The p-values are produced by the `Detector` module in the drift detection pipeline using a BBSE (trained LSTM model) for the Reductor and MMD test for the Tester.

**Figure 6:**
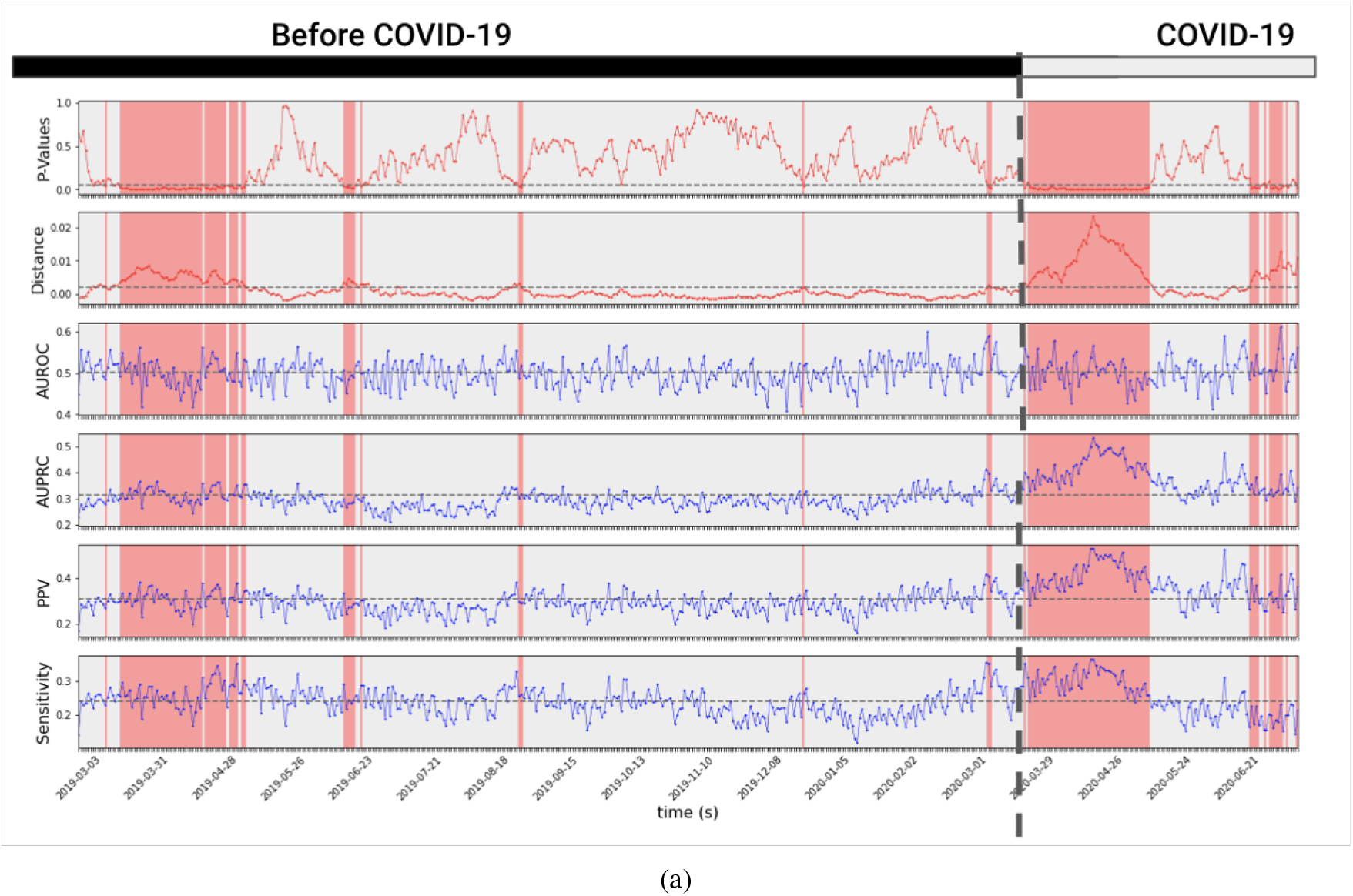
Monitoring data shifts in mortality decompensation prediction from 03/2019 to 07/2020 using a 30-day `RollingWindow`. It can be seen that drift is detected in 03/2020 (red), which corresponds to when Ontario declared the COVID-19 pandemic a state of emergency and entered a province-wide lockdown.

### 5.2 Medical Information Mart for Intensive Care (MIMIC)-IV

MIMIC-IV is a single-center database comprising information relating to a diverse and very large population of ICU patients between 2001 and 2012. The database includes static (e.g., demographics and baseline measures) and temporal information (e.g.,vitals, medications, and interventions) extracted from several sources including archives from critical care information systems, hospital EMR, and Social Security Administration Death Master File. MIMIC-IV is a relational database consisting of 26 tables and is freely available to researchers worldwide.

#### 5.2.1 Mortality Decompensation Prediction

We used the MIMIC-IV database to demonstrate the usability of CyclOps framework for mortality decompensation prediction task among critical care patients. Details of the queried data, process API parameters, and the vectorized data are shown in Table 1. Modeling was performed with the same setting as in mortality decompensation prediction for GIM patients, however we made use of chart events which includes observational data obtained both from GIM as well ICU environments. The LSTM model was trained on 53,386 records and evaluated on 6,673 records with an AUROC score of 0.86.

### 5.3 NIH Chest X-Ray Dataset

We utilized the chest X-ray dataset from NIH [23] to demonstrate the usability of CyclOps framework for detecting distribution shift in medical imaging data. The dataset contains 112,120 frontal-view X-ray images of 30,805 unique patients with fourteen disease image labels extracted from the corresponding text-based radiology reports. The creators of the dataset have already split the data into a training set of 80,000 images, a validation set of 10,000 images, and a test set of 10,000 images. Using the drift detection pipeline, we performed two synthetic shift experiments where the images in the test set were synthetically perturbed using the `SyntheticShiftApplicator`. In our first experiment, we corrupted the samples in the test set with varying levels of Gaussian noise which formed our target dataset. In our second experiment, we performed categorical shifts by sub-sampling the target dataset based on patients’ demographics information (age, gender, or ethnicity). This would artificially change the distribution of patients’ demographics across observed classes. For both experiments, we initialized the `Reductor` with a pre-trained autoencoder from the TorchXrayVision library [1] to reduce the dimensions of the images, and MMD as for the Tester. Using the `Detector` we configured with the `Reductor` and `TSTester`, we performed sensitivity tests and obtained the p-values shown in Figure 7.

**Figure 7:**
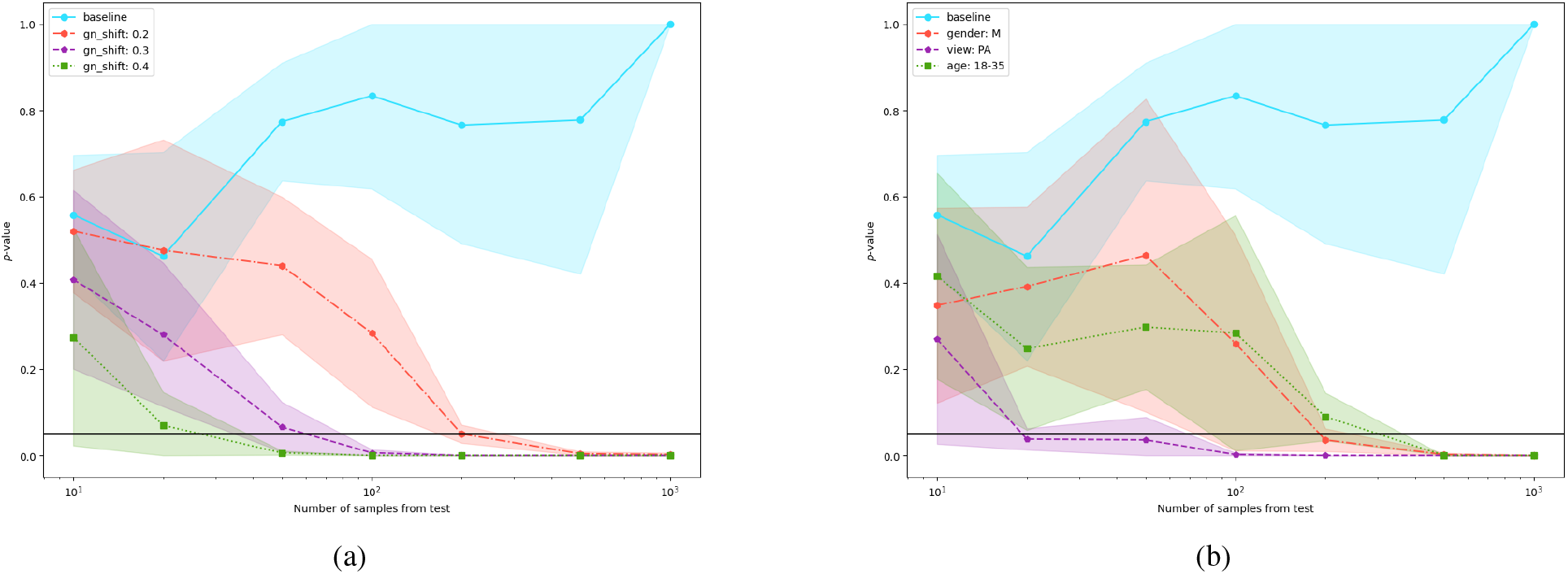
Drift Detection experiments on NIH Chest X-Ray data. a) Gaussian noise injection experiments for an increasing amount of Gaussian noise. b) Categorical shift experiment to simulate population shifts based on categorical metadata.

## 6 Conclusion

In this paper, we introduced the CyclOps framework, its design principles, and its usability across various use cases and datasets at the intersection of ML and health. CyclOps is an extensible, modifiable, and reusable framework which was designed to facilitate the adoption of ML models at scale and enable building sophisticated ML models on large volumes of data. More importantly, CyclOps was intended to empower operationalizing ML workflows through its cyclical development consisting of data, model development and monitoring pipelines.

## Data Availability

All data produced in the present study are available upon reasonable request to the authors, subject to governance policies of datasets used for the experiments.

https://www.geminimedicine.ca/

https://physionet.org/content/mimiciv/2.1/

## 7 Acknowledgements

This work is made possible due to the data obtained from the General Medicine Inpatient Initiative (GEMINI), and we acknowledge the GEMINI team for their support. We also acknowledge HPC4Health, for enabling high performance computing environments which were involved in the development of this work. Finally, we acknowledge support from the Vector Institute and its vibrant community working at the intersection of health and machine learning.

